# Subtle anomaly detection in MRI brain scans: Application to biomarkers extraction in patients with *de novo* Parkinson’s disease

**DOI:** 10.1101/2021.06.03.21258269

**Authors:** Verónica Muñoz-Ramírez, Virgilio Kmetzsch, Florence Forbes, Sara Meoni, Elena Moro, Michel Dojat

**Affiliations:** Univ. Grenoble Alpes, Inria, CNRS, Grenoble INP, LJK, 38000 Grenoble, France; Univ. Grenoble Alpes, Inserm U1216, CHU Grenoble Alpes, Grenoble Institut des Neurosciences, 38000 Grenoble, France

**Keywords:** Autoencoder, Machine learning, Neurodegenerative Disease, Parkinson’s Disease, Variational autoencoder

## Abstract

With the advent of recent deep learning techniques, computerized methods for automatic lesion segmentation have reached performances comparable to those of medical practitioners. However, little attention has been paid to the detection of subtle physiological changes caused by evolutive pathologies such as neurodegenerative diseases. In this work, we investigated the ability of deep learning models to detect anomalies in magnetic resonance imaging (MRI) brain scans of recently diagnosed and untreated (*de novo*) patients with Parkinson’s disease (PD). We evaluated two families of auto-encoders, fully convolutional and variational auto-encoders. The models were trained with diffusion tensor imaging (DTI) parameter maps of healthy controls. Then, reconstruction errors computed by the models in different brain regions allowed to classify controls and patients with ROC AUC up to 0.81. Moreover, the white matter and the subcortical structures, particularly the substantia nigra, were identified as the regions the most impacted by the disease, in accordance with the physio-pathology of PD. Our results suggest that deep learning-based anomaly detection models, even trained on a moderate number of images, are promising tools for extracting robust neuroimaging biomarkers of PD. Interestingly, such models can be seamlessly extended with additional quantitative MRI parameters and could provide new knowledge about the physio-pathology of neuro-degenerative diseases.

## 1. Introduction

Deep Learning (DL) is a growing trend in image analysis. Recent breakthroughs, notably the explosion of the available computer power and the availability of large datasets, have allowed artificial neural networks to obtain state-of-the art performances in several computer vision challenges (i.e. ILSVRC2012 http://image-net.org). These developments have also been exploited successfully in several medical applications (Esteva et al., 2017; Poplin et al., 2018; Titano et al., 2018). A particular interest in DL methods has raised with the hope to create support tools for radiologists to analyze images, segment lesions and detect subtle pathological changes that even an expert eye can miss (Hosny et al., 2018; Topol, 2019; Bernal et al., 2019).

DL methods are hungry data consumers. Over the years, the MRI community has curated several public databases with annotated images to test lesion segmentation algorithms. Recently DL approaches have defeated all alternative methods in challenges such as MSSEG, for multiple sclerosis lesion segmentation (http://portal.fli-iam.irisa.fr/msseg-challenge); BRATS, for brain tumor segmentation (www.braintumorsegmentation.org); ISLES, for ischemic stroke lesion segmentation (www.isles-challenge.org); and mTOP for mild traumatic injury outcome prediction (www.tbichallenge.wordpress.com). Moreover, the recent development of unsupervised anomaly detection techniques has opened the door to many more applications of lesion identification, specially for pathologies where annotated data is scarce and other, like Parkinson’s Disease (PD), where lesions are not visible to the naked eye.

Although they may vary in architecture and loss function, unsupervised methods often follow the same steps. First, they are trained exclusively with images from the “normal” class. Second, during testing, they are exposed to images that contain anomalies for which they cannot provide an accurate reconstruction. Finally, the resulting reconstruction errors are leveraged to locate extreme values, considered as anomalies. Auto-encoders (AE) and deep generative models, such as variational auto-encoders (VAE) (Kingma and Welling, 2019) and generative adversarial networks (GAN) (Goodfellow et al., 2014), have been extensively used as building blocks for unsupervised anomaly detection due to their ability to learn highdimensional distributions. In this paper, we designed a pipeline, using AEs and VAEs at its core, for extracting biomarkers in magnetic resonance imaging (MRI) brain scans of PD patients.

### 1.1. Medical Context

PD is the second most prominent neurodegenerative disease in the world, affecting more than six million people worldwide. Symptoms generally develop slowly over the years. During the podromal phase, the loss of dopamine-producing neurons disrupts the functioning of several subcortical structures. Several non-motor symptoms appear such as visual and olfactory disturbances or mood disorders. However, generally, the diagnosis occurs once the patients start experiencing the well-known motor symptoms of PD, namely stiffness, akinesia and resting tremor. By this moment, it is estimated that more than 60% of dopaminergic neurons have already been lost or impaired.

The manifestation of non-motor symptoms (i.e. olfactory dysfunction, sleeping troubles, autonomic disturbances) during the prodromal stage of PD indicates the presence of physiological changes that could be used to characterize PD patients. Indeed, large research efforts are currently performed to find biomarkers that allow earlier diagnosis and patient sub-typing to improve treatment (Barber et al., 2017; Peran et al., 2010).

MRI has the potential to reveal robust neuroimaging biomarkers for the disease progression monitoring and long-term drug impact analysis. As a matter of fact, MRI is not only useful to observe qualitative changes in the structural integrity of the brain but is also a valuable tool to characterize quantitative physiological features such as blood perfusion and water diffusion. Diffusion MRI monitors the displacement of water molecules in the brain. As their rate and direction of movement are dependent on their environment, diffusion measurements give indirect information on the structural composition of the brain (Le Bihan and Johansen-Berg (2012)). In order to better understand the progression of the disease, we used diffusion MRI to study the undergoing brain changes in *de novo* PD patients (i.e. newly diagnosed and without dopaminergic treatment) compared to healthy controls (HC).

### 1.2. Contributions

We propose the use of fully convolutional autoencoders and variational auto-encoders to provide a meaningful representation of MRI data from healthy brains. Once these models are trained, they are used to identify unusual patterns in data from *de novo* PD patients. This is not without challenge. In contrast to other medical imaging applications, such as the segmentation of brain tumors or MS lesions, we do not dispose of a ground truth that identifies, at the voxel level, values specific to PD, especially for *de novo* patients. This complicates model evaluation and selection. Indeed, the only available information is the classification of each individual as PD patient or HC. In addition, PD-related abnormalities may be very local and particularly subtle to detect in *de novo* patients. Indeed, at this stage PD affects only a few neighboring voxels, mainly located in subcortical structures, such as the Substantia Nigra (SN), as opposed to larger brain lesions such as brain tumors, stroke, or T2-hyperintense lesions in MS, clearly visible in MRI. A last, to eliminate additional sources of bias, it is wise to consider scans with homogeneous acquisition parameters, notably the same magnetic field and scanner manufacturer. As a consequence most medical studies tend to provide training data of reasonable size in a clinical context but limited with respect to the usual deep learning requirement. The neuroimaging database available for this study contains MRI volumes from 56 healthy controls (see section 3.1).

For all these reasons, the task of finding subtle anomalies in PD brain scans is not the most straightforward for DL techniques. Despite the rising interest in uncovering biomarkers of PD and the advent of DL for anomaly detection, no studies have been published, to the best of our knowledge, concerning DL approaches for PD diffusion alterations discovery. The main contribution of our work is then two-fold:

1. We demonstrate the appropriateness of fully convolutional auto-encoders and variational auto-encoder architectures, which preserve spatial information in the latent space, for anomaly detection in MRI data of recently diagnosed PD patients. Although only two diffusion MRI measures were used for this study, the presented approach is extensible to explore the predicting power of additional parameters.
2. We propose a performance evaluation method that may be used to analyze neuroimaging anomalies in other disorders where no voxel-level ground truth is available.

This paper is organized as follows. Section 2 reviews state-of-the-art approaches to detect and characterize abnormal regions in MR brain scans of PD patients, as well as examines recently published DL frameworks for anomaly detection in other medical applications. Section 3 describes the datasets we used and the implemented models. The experimental setup and procedures are described in Section 4 while the obtained results are featured in Section 5. Finally, Section 6 explores the significance of our results and Section 7 summarizes the impact of this study and gives directions for future work.

## 2. Related Work

### 2.1. Diffusion MRI for the study of Parkinson’s disease

Diffusion changes in PD has been the subject of many studies over the years. They are generally based on two measures accounting for mean diffusivity (MD) and fractional anisotropy (FA). These measures describe the diffusion of water molecules in the brain, MD accounts for the their overall displacement and FA indicates the orientation of diffusion. The meta-analysis proposed by Cochrane and Ebmeier, 2013 put into evidence important discrepancies between the diffusion scans of PD patients across studies between 1946 and 2012, notably regarding acquisition parameters, analysis methods and the introduction of medication. Most studies focused on the SN as Region Of Interest (ROI) and often reported FA reductions in different segments with a slight tendency towards the caudal segment. However, no significant association was detected between disease severity and FA values. The first studies were carried out on small cohorts (around 50 individuals) and presented opposing results. We can notably cite the work of Schwarz et al., 2013 where, in contrast to the work of Du et al., 2011, no differences were found in SN for FA values between PD patients and controls but a significant increase of MD in the SN (P < 0.005) was reported.

A longitudinal study that follows *de novo* PD patients of 35 centers for five years has been carried out by the Parkinson’s Progression Markers Initiative (PPMI) (Marek et al., 2018). The corresponding database is openly available for researchers and contains, among other clinical and behavioral assessments, MR structural and diffusion imaging scans. Based on PPMI data, in a large study of SN abnormalities, Schuff et al. (2015) consider the laterality of the pathology. A significant FA reduction (p=0.04) in PD patients was found in the rostral part of the SN contralateral to the body side presenting the greater functional deficits. Furthermore, significant relationships were reported between these results and the dopaminergic deficits displayed in dopamine transporter (DAT) scan images. Nevertheless, contralateral rostral FA values allowed to only achieve a classification ROC AUC of 59% using a bootstrapped half-split cross-validation procedure.

The meta-analysis presented by Atkinson-Clement et al. (2017) explored DTI alterations beyond the SN. The disease effect size (*D*_*ES*_) of FA and MD were analyzed in 27 anatomical ROIs. White matter fiber degradation in PD was associated to reduced values of FA-*D*_*ES*_ and increased MD-*D*_*ES*_ in the SN, the corpus callosum, the cingular and temporal cortices. In contrast, the corticospinal tract displayed increased FA-*D*_*ES*_ and decreased MD-*D*_*ES*_ .

In the work of Cousineau et al. (2017), 50 white matter tracts connecting subcortical structures of the brain to different cortical areas were dissected. Welch’s unequal variance t-test was employed to identify the sections of those tract profiles that were significantly different between PD patients and controls from the PPMI. The analysis found statistically significant differences in tract profiles along the subcortico-cortical pathways between PD patients and healthy controls. In particular, significant increases in FA, apparent fiber density and tract-density were detected in some locations of the SN-STN-PutamenThalamus-Cortex pathway, which is one of the major motor circuits driving the coordination of motor output.

Up until this point, the studies on PD diffusion alterations suffer from great heterogeneity. Some are carried out with a small number of participants and others are meta-analysis. They present inconsistent findings about the spatial location of brain abnormalities which raise questions as to whether standard statistical methods are capable of bringing out robust biomarkers for such a complex disease. Recently data mining classification tools such as support vector machines (SVM) have been applied to the identification of PD abnormalities. Talai et al., 2018 employed them to differentiate PD from other PD syndromes such as progressive supranuclear palsy (PSP). Using a feature selection method, the FA and MD average values of 17 ROIs were chosen to train a SVM in a leave-one-out cross-validation procedure. The classifier was able to differentiate PD and PSP patients with an accuracy of 87.7%. The main affected regions were the brainstem, putamen, palladium, thalamus and some areas of the frontal cortex. In a similar study, Correia et al. (2020) went farther and proposed to employ SVM to classify between controls, PD, PSP and corticobasal syndrome (CBS) patients with FA and MD measures in selected regions of white matter. The mean accuracy for the leave-two-out cross-validation to separate controls from PD patients was of 61.3%.

To conclude, different studies (Du et al., 2011; Schwarz et al., 2013; Atkinson-Clement et al., 2017; Schuff et al., 2015) have studied diffusion changes in PD patients. Most of them exploit standard statistical methods, such as twosample t-tests, to analyze voxel-wise differences between MRI data from PD patients and healthy controls. However, inconsistent findings, as shown by Schwarz et al. (2013) and Atkinson-Clement et al. (2017), suggest that novel approaches are required to truly take advantage of the rich information captured by diffusion MRI and discover powerful biomarkers for such a complex neurodegenerative disease.

### 2.2. Deep learning for anomaly detection in medical images

Numerous techniques have been proposed for anomaly detection using artificial neural networks (see Chalapathy and Chawla (2019) for a recent review). Most state-ofthe art DL anomaly detection techniques use Generative Adversarial Networks (GAN) or Auto-Encoders (AE) and their variations. One of the precursor works of anomaly detection applied to medical data was proposed by Schlegl et al. (2017) to identify anomalous regions in spectral-domain OCT scans of the retina. Their architecture, named AnoGAN, consisted on a convolutional GAN architecture trained with 2D patches that achieved a ROCAUC of 89%. This work demonstrated that unsupervised learning on large-scale imaging data could lead to the detection of relevant anomalies. As presented by Alex et al. (2017), another possibility is to use the discriminator of the trained GAN architecture to output, for each query image, a probability map that gives an indication of the likelihood for every point of belonging to the learnt “normal” data distribution. The authors applied the proposed method to detect brain tumors from the multimodal MR images (FLAIR, T2, T1, T1 post contrast) of the BRATS dataset. The model was trained with 2D patches and obtained a DICE score of 69 % for the whole tumor segmentation task. Recently, Ha Son et al. (2019) proposed to increase the training stability of GANs by employing two auto-encoders as generator and discriminator. Their model, called ADAE (Adversarial Dual Auto-encoders), was also trained on BRATS data and achieved a ROC AUC score of 89.2%.

In contrast to models based on the reconstruction error, Zimmerer et al. (2018) claimed that a Variational AutoEncoder (VAE) gradient-based *score* approximation is better suited for anomaly detection. The proposed method assigns an anomaly rating to each pixel: the derivative of the log-density with respect to the input, evaluated through backpropagation. These two frameworks leverage the available voxel-level ground truth and use strong post-processing techniques to smooth the output. In addition, they do not deal with image resolution higher than 64 × 64 pixels. For these reasons, there is considerable uncertainty with regard to their usefulness when no ground truth is available at the voxel level.

The first DL-based anomaly detection approach that operates on entire brain MRI slices (instead of patches) was proposed by Baur et al. (2019). The authors introduce *AnoVAEGAN*, combining concepts of VAE and GAN and alternating between two loss functions: one for the generator (VAE component aiming at faithfully reconstructing the inputs) and one for the discriminator (to distinguish between real and reconstructed MRI). To evaluate the model, the authors trained the model with healthy FLAIR scans and attempted to detect MS lesions. They compared their results to those obtained by the precursor AnoGAN (Schlegl et al., 2017) and by dense and spatial variants of VAEs and AEs. DICE scores showed that AnoVAEGAN beat AnoGAN (0.605 vs. 0.375) as well as dense and spatial variations of VAEs and AEs. However, spatial VAE (0.592) and spatial AE (0.585) performed only slightly worse than their model without any adversarial training. *AnoVAEGAN* demonstrated that DL-based anomaly detection worked in fairly high-resolution images, and that abnormal regions could be identified by pixel-wise intensity difference between the input image and its reconstruction. Moreover, it showed that simple models (regular auto-encoders and VAE) may be used for anomaly detection with only marginally inferior results than more complex models, as long as spatial information is maintained in the latent space. While a DICE score of 60.5% may seem poor compared to the previous DL results here exposed, MS lesions are generally more challenging to segment than tumors (Commowick et al., 2018).

Using such an anomaly detection approach for the investigation of alterations in the brain of PD patients constitutes a great challenge as early symptoms in PD have not yet been translated to identifiable characteristics in MR imaging. Regardless, in the last years some innovative methods have been devised to apply unsupervised deep learning to PD anomaly detection in MR imaging. Li et al. (2019) set out to employ a stacked sparse autoencoder (SSAE) to discriminate between controls and PD patients from longitudinal data pooled from the PPMI database. T1 and DTI maps were collected for every subject at baseline, after 12 months and over 24 months. Gray matter, white matter and mean diffusivity features were extracted for every one of the 116 ROIs in the AAL2 atlas and serve as input for the SSAE. The outputs of the architecture at the three time-points were classified with an SVM. The proposed method achieved a ROC AUC score of 86% at baseline and 97% at 24 months. They compared their results with a simple sAE, a CNN and a DBN architecture. The sAE achieved the second best results with a ROC AUC of 82% at baseline and 92% at 24 months. Other studies preferred to employ CNNs to discriminate *de novo* PD patients from HC scans. Shinde et al. (2019) adapted the well know ResNet50 design to study 2D NMS-MRI images of the SN from a local database. They achieved a good classification performance (ROC-AUC = 90%) taking into account clinical laterality. Sivaran-jini and Sujatha (2020) proposed to employ the popular AlexNet CNN to discriminate T2-weighted MR scans of HC and PD patients from the PPMI at various stages of the disease. One particularity of this investigation is the utilization of transfer learning, that is, the CNN architecture had been pre-trained with natural images before medical images were input. Their results show an accuracy of 88.9% corresponding to 89.3% in sensitivity and 88.4% in specificity.

In summary, few deep learning studies have ventured in the search of PD anomalies. The study presented in this paper covers an uncharted territory by studying brain abnormalities in *de novo* and untreated PD patients with reconstruction methods.

## 3. Materials and methods

### 3.1. Data description

The dataset used in this work consists of DTI MR scans coming from the PPMI database. DTI data are particularly sensitive to various sources of bias (Jones and Cercignani, 2010). To moderate this effect, we selected DTI scans from 56 healthy controls and 129 *de novo* PD patients acquired with the same MR scanner model (3T Siemens Trio Tim) and configured with the same acquisition parameters. From these images, two measures, mean diffusivity (MD) and fractional anisotropy (FA), per voxel were computed using MRtrix3.0. Values of FA and MD were normalized into the range [0, 1]. Each MRI volume was obtained in the NIfTI format as a 3dimensional, 121 × 145 ×121, array with a voxel size of 1.5 × 1.5 × 1.5mm^3^ . Only 21% ± 1.9% of this volume was effectively occupied by the brain and cropped images of 40 axial slices around the center of the brain were used to train the models. In addition, we considered the right and the left hemispheres images as two different samples. The left hemisphere images were flipped to resemble the right hemisphere images. This increased the number of images available for training and allowed us to study the laterality of PD. Hemispheres were preferred over smaller regions to preserve spatial information as advised in the work of Baur et al. (2019), which demonstrated the usefulness of spatial AE and VAE models for anomaly detection in high-resolution MRI. Finally, all 80 individual axial hemisphere slices were cropped to a 75 × 145 resolution keeping the brain at the center of the images and thus ensuring consistency for our models. The control dataset was divided into 41 training controls and 15 testing controls to avoid data leakage. As a result the models were trained with a duo (FA & MD) of 1680 images (2 hemispheres × 40 slices × 41 controls).

As stated in Poldrack et al. (2019), it is dangerous to infer predictive accuracy based on a model that is constructed for a specific population. To assess the influence of the training and test populations on the final results, a bootstrap procedure was carried out. We divided the control dataset into training and testing samples in 10 different manners, taking special care to maintain an age average around 61 years old for all the training and test population as well as a 40-60 proportion of females and males. Table 1 shows the age and gender characteristics of the training and test sets of the ten sub-populations.

**Table 1:** Training and test age distributions for 10 control populations

|  | Training set | Testing set |
| --- | --- | --- |
| Sample 1 | $60.9 \pm 10.5$ | $61.5 \pm 8.2$ |
| Sample 2 | $61.3 \pm 9.2$ | $60.7 \pm 11.9$ |
| Sample 3 | $61.2 \pm 10.2$ | $60.9 \pm 9.3$ |
| Sample 4 | $61.0 \pm 10.0$ | $61.3 \pm 9.8$ |
| Sample 5 | $61.1 \pm 9.5$ | $61.1 \pm 11.2$ |
| Sample 6 | $61.1 \pm 9.5$ | $61.0 \pm 11.2$ |
| Sample 7 | $61.1 \pm 10.2$ | $61.1 \pm 9.2$ |
| Sample 8 | $61.1 \pm 10.6$ | $61.1 \pm 7.8$ |
| Sample 9 | $61.3 \pm 10.1$ | $60.4 \pm 9.5$ |
| Sample 10 | $61.1 \pm 10.3$ | $61.1 \pm 9.0$ |

Once the models were trained with one of the 10 training sets, they were evaluated with the corresponding healthy control test set and the PD dataset.

### 3.2. Architecture design

We chose to introduce multiple MRI parameter maps as input in a joint manner, as evoked in the work of Baur et al. (2019). Our inputs were of dimension *H* × *W* × *C*, where *H* represents the height in pixels, *W* the width in pixels and *C* the number of channels (MRI measurements). For the PPMI data used in this work, *H* = 145, *W* = 75 and *C* = 2. Our approach is straightforwardly adaptable to leverage the predicting capability of more MRI parameters by concatenating more channels in the input images.

The anomaly detection task with an auto-encoder can be formally posed as follows. We consider an input image *x* ∈ ℝ ^*H*×*W*×*C*^ and a trained auto-encoder with encoder *f*_φ_ and decoder *g*_θ_. The encoder maps the input *x* to a lower dimensional latent representation *z*, then the decoder maps the latent vector *z* to the reconstructed output 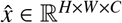:

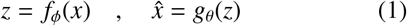

The pixel-wise reconstruction errors are computed as 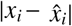 Depending on the network architecture, the latent code may be a simple vector (*z* ∈ ℝ ^*d*^) or a third-order tensor (*z* ∈ ℝ ^*h*×*w*×*c*^). The former is referred as a dense bottleneck and the latter as a spatial bottleneck.

For a VAE, the difference is that the encoder generates the parameters of the approximate posterior of the latent variable given the input, constrained to follow a multivariate normal distribution, and a sampling operation is needed to obtain an actual value for *z*:

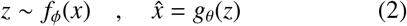

We constructed and evaluated three autoencoder-based models: a spatial variational auto-encoder (sVAE), a spatial auto-encoder (sAE) and a dense variational autoencoder (dVAE).

All models were implemented using Python 3.6.8, PyTorch 1.0.1, CUDA 10.0.130 and trained on a NVIDIA GeForce RTX 2080 Ti GPU with batches of 32 images. After each convolutional layer, batch normalization (Ioffe and Szegedy, 2015) was applied for its regularization properties and to avoid vanishing or exploding gradients. The nonlinear activation function in each layer except the last was the rectified linear unit (ReLU), which was defined as *f* (*x*) = *max*(0, *x*). In the last layer, the activation function was a sigmoid, defined as *f* (*x*) = 1/(1 + *e*^−*x*^), in order to have output pixels normalized between [0, 1]. The loss functions were optimized using Adam (Kingma and Ba, 2015), a popular optimization algorithm for training deep neural networks. The differences between each of the four models are detailed below.

#### 3.2.1. Spatial auto-encoder

The spatial auto-encoder model was fully convolutional, 5 convolutional layers connected inputs to bottleneck and 5 transposed convolutional layers from bottleneck to outputs. As depicted in Figure 1-B, the output of the encoder network was directly the latent vector *z* and the loss function was simply the *L*_1_-norm reconstruction error:

**Figure 1:**
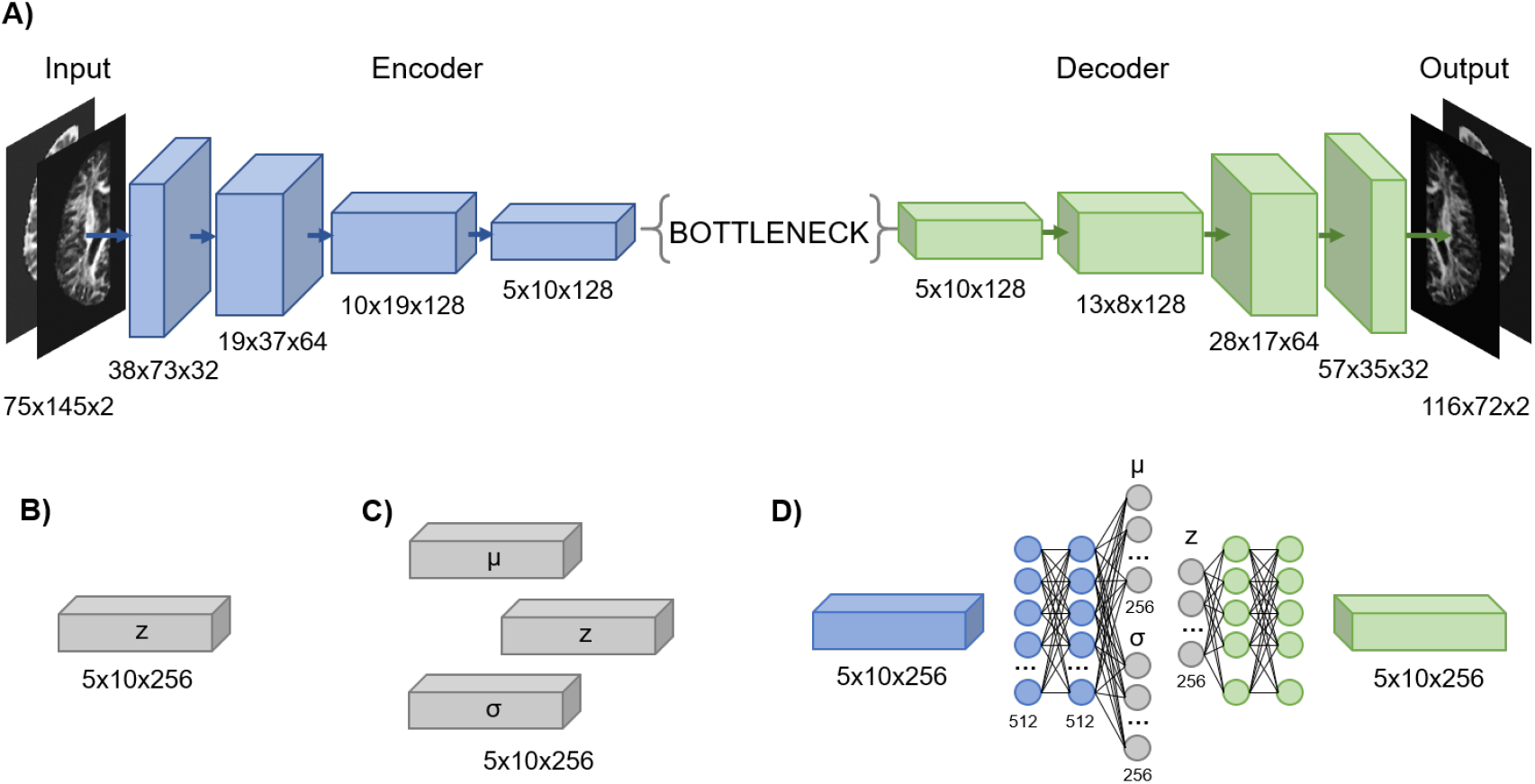
A) General architecture of the implemented auto-encoder with an unspecified bottleneck; B) sAE spatial bottleneck; C) sVAE spatial bottleneck and D) convolutional layer of the dVAE along with its fully connected layers and dense bottleneck. *µ* and *σ* describe the approximate posterior of the latent variable, *z* was obtained by a sampling operation.

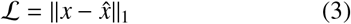

This model was trained for 160 epochs, with a learning rate of 0.8 × 10^−3^. 3 × 3 kernels were convoluted using padding of 1 pixel and a stride of (2, 2) in the first four layers and a stride of (1,1) in the last one. The bottleneck dimensions were h=10, w=5 and c=256. There were no pooling layers.

#### 3.2.2. Spatial variational auto-encoder

Similar to the sAE, the model was fully convolutional, however, the encoder generated the parameters of the approximate posterior of the latent variable given the input, constrained to follow a multivariate normal distribution. A sampling operation was needed to obtain an actual value for *z* as depicted in Figure 1-C.

Training lasted for 200 epochs using a learning rate of 0.3 × 10^−3^. As in the previously presented model, a 3 × 3 kernel was chosen as filter, along with a padding of 1 and stride of (2, 2) in the first four layers and of (1,1) in the last one, resulting in bottleneck dimensions h=10, w=5 and c=256. No pooling layers were used. The loss function was computed as follows:

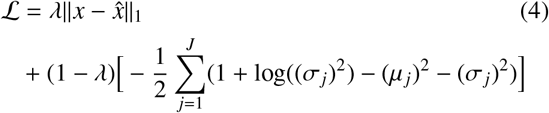

where *µ* and *σ* denote the mean and the variance of the approximate posterior, *J* is the number of elements in the latent space and λ controls the proportion between the two terms. The first term is the reconstruction error and the second term is the Kullback-Leibler (KL) divergence between the approximate posterior and the prior of the latent variable, for the Gaussian case (Kingma and Welling, 2019). To favor good reconstructions over a Gaussian-like distribution of the latent variables, we put more weight (90%) in the reconstruction term and less weight (10%) in the KL divergence term. This proportion was empirically determined to reduce reconstruction error while improving anomaly detection.

#### 3.2.3. Dense variational auto-encoder

The main difference of the dense variational autoencoder when compared to the sVAE is its dense bottleneck. Encoder and decoder also have fully connected layers in addition to the convolutional layers, as shown in Figure 1-D. The number of units is depicted below each fully connected layer. The dimensions of the outputs of each convolutional layer (not shown in Figure 1) were the same as for the sVAE model.

For regularization purposes, the dropout technique (Srivastava et al., 2014) was used to turn off 30% of the units in fully-connected layers during training. This model was trained for 100 epochs with a learning rate of 0.3 × 10^−3^. There were no pooling layers. Kernels for all convolutional layers were 3 × 3, and convolutions were performed with a padding of 1 and stride of (2, 2). The bottleneck vector was of size d=256.

The same modification in the VAE loss function was implemented in this model, as shown in Equation 4, and we kept the same 90/10 proportion between the reconstruction term and the KL divergence term.

## 4. Evaluation procedure

After training, the models were fed with the images from the test controls and PD patients datasets. As already mentioned (Table 1), this was done 10 times using different training sets.

The idea was to identify diffusion anomalies in PD patients by observing for which brain regions the models provided an inaccurate reconstruction while providing accurate ones for healthy test controls. We expected the reconstructions of healthy subjects be superior to those of patients because the models were trained solely with healthy scans. That being said, we also expected to reveal, if any, subtle anomalies since all patients were only recently diagnosed. For this purpose we computed a joint reconstruction error map for all outputs where the value for each voxel *i* was calculated as:

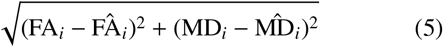

FA_*i*_ and MD_*i*_ denote the original FA and MD values while 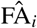 and 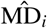 the reconstructed values.

We could infer that reconstruction errors come from at least four different sources: 1) noise in the input data, 2) loss of information due to dimension reduction in the latent space, 3) variability of healthy controls not captured by the model and 4) finally real anomalies caused by PD. Because we were only interested in the latter, the best way to evaluate and compare the models was by measuring their ability to discriminate between controls and PD patients, based on the intensity and localization of the reconstruction errors.

### 4.1. Regions of Interest

In order to evaluate the reconstruction of specific ROIs, we used two brain atlases. The Neuromorphometrics atlas (Bakker et al., 2015) was used to localize the Frontal, Temporal, Occipital and Parietal lobes, the Cingulate and Insular cortex and the overall White Matter. The MNI PD25 atlas (Xiao et al., 2015), specifically designed for PD patients MRI analysis, was used to localize several subcortical structures, the Red Nucleus (RN), the Substantia Nigra (SN), the Subthalamic Nucleus (STN), the Caudate Nucleus, the Putamen, the external and internal Globus Pallidus and the Thalamus.

### 4.2. Anomaly identification

In order to detect abnormal voxels in subjects, we needed to decide above which error value (hereafter the *abnormality threshold (a*.*t*.)), a given voxel was considered as poorly reconstructed. While large error values could also occur for voxels in healthy subjects, they should appear much less frequently than for PD patients as the later were not used to train the network. Therefore, the *a*.*t*. value was fixed to an extreme quantile (*e*.*g*. the 98% quantile) of the distribution of errors in the control population. This meant that only a very small percentage (2%) of the voxels in the control population were classified as abnormal due to subject variability and learning imperfections. Next, for each control and PD subject in the test set, we counted the number of abnormal voxels detected in each ROI as a percentage. A subject was then classified as PD if this percentage was above a certain value that had to be decided. The classification performance depends on this choice. In practice, we proposed a ROC curve analysis to set a reasonable value of this threshold referred to as *pathological threshold (p*.*t*.).

Due to the imbalanced nature of the test set (30 healthy and 258 pathological brain hemispheres), considerable care was taken to choose a reliable criterion for classification performance evaluation. The trade-off between sensitivity and specificity depicted by the ROC curve deemed itself the best adapted as it takes into account the ability to identify both classes independently from class prevalence. The area under the ROC curve (AUC) can be directly used as a measure of classification performance. In addition, as each point in the ROC curve corresponds to the sensitivity and specificity values obtained for a given threshold, the point closest to a perfect sensitivity and specificity can be considered as the *p*.*t*. This is illustrated on Figure 2. And so, the *abnormality threshold* serves for classification at the voxel-level, while the *pathological threshold* differentiates subjects at the hemisphere-level or ROI-level. The classification performance of the pathological threshold can be evaluated using the geometric mean (g-mean) between sensitivity and specificity :

**Figure 2:**
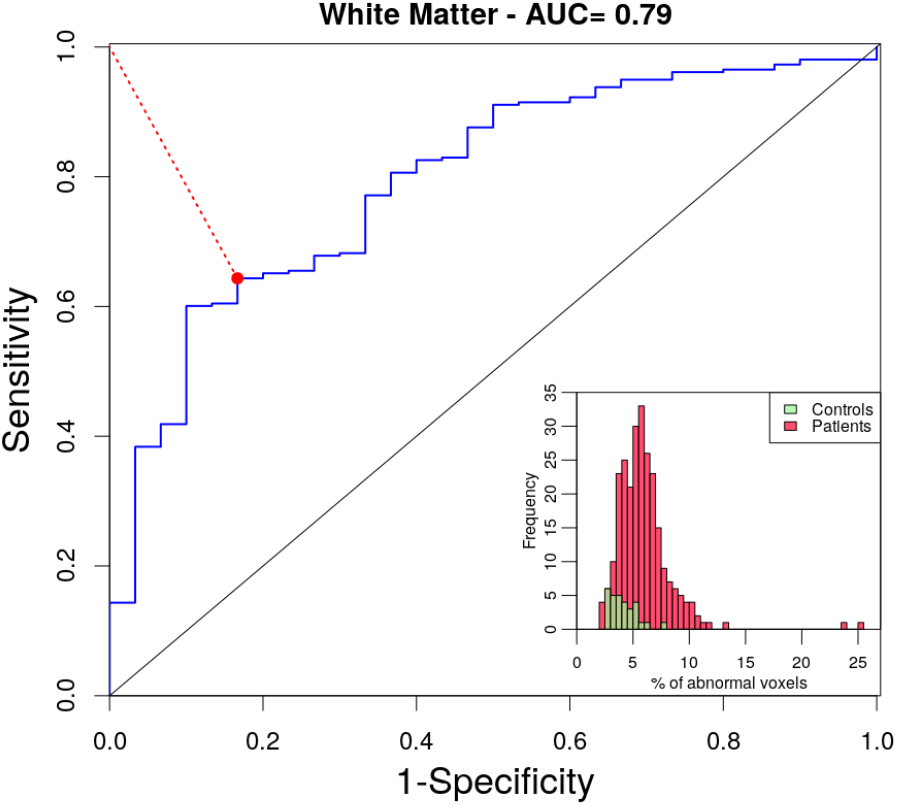
ROC curve obtained when varying the percentage of abnormal voxels in white matter (WM) above which a subject is classified as PD. Sensitivity and specificity are computed using controls and patients reconstructions. The retained threshold for PD patient vs Control classification correspond to the red point. Insert: Histogram of WM abnormal voxels percentages.

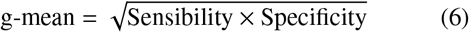

For all of the before-mentioned ROI, we calculated the ROC-AUC and the g-mean of the associated pathological threshold. The results across all 10 sub-populations for all models are reported in Section 5.1.

We also investigated the classification performances obtained with different sub-types of patients depending on their clinical history, mainly concentrating on cognitive compliance and motor symptoms. Some studies have found that at the early stages of PD, anomalies were often detected in patients that present mild cognitive impairment (MCI) versus patients without cognitive symptoms (Tessa et al., 2014; Zeighami et al., 2017; Malek et al., 2015; Guimarães et al., 2018). To test for this possibility in our dataset, we divided the patients into two categories based on their MoCA test scores that reflect their cognitive skills. The patients with a score below 26 points were classified as MCI. PD patients often present unilateral motor symptoms at the early stages of the disease. In this case, we expect a brain alteration in the contralateral hemisphere. Inspired by other studies (Schuff et al., 2015; Shinde et al., 2019) we tested the prediction differences between hemispheres based on the UPDRS III test scores, which reflect motor dysfunctions. An hemisphere was considered as altered when the sum of the UPDRS III scores related to the contralateral motor side was equal or above 10 points.

## 5. Results analysis

As it can be observed in Figure 3, the best reconstructions were achieved by the sAE. This was expected as there were no regularization constraints for the reconstructions. The sVAE generated quite good reconstructions as well. On the contrary, the dVAE was unable to reconstruct fine details in the images, specially around the circonvolutions and the subcortical structures. This may be caused by the loss of information in its fully connected layers. Supervised learning applications of auto-encoders in the literature have also shown that dVAE are very sensitive to outliers and often struggle to reconstruct detailed structures in brain scans (Baur et al., 2019). However, due to the lack of ground truth, we cannot conclude on which model provides the better performances.

**Figure 3:**
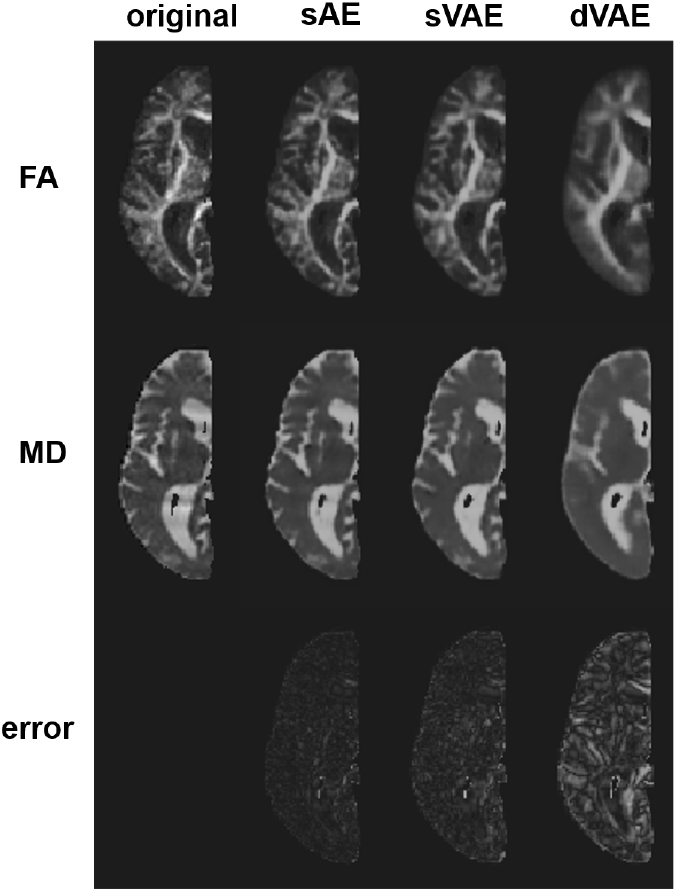
Original FA (top) and MD (middle) axial brain slices of a PD patient with their corresponding reconstructions using the sAE, sVAE and dVAE models. The joint reconstruction error maps are displayed in the bottom row.

### 5.1. Most discriminant ROIs

It would be beneficial to find a structure which, when abnormal, is both PD specific and sensitive enough to discriminate most *de novo* PD patients from HC. The first performance evaluation corresponds to the ROC-AUC obtained when comparing, for each ROI and each subpopulation, the percentage of values above the 98% quantile of the reconstruction error distribution for HC. The results displayed in Figure 4 show that comparing the percentages of abnormal voxels in the whole brain yields satisfactory classifications with an average AUC of 67.6%, 67.5% and 63% for the sAE, sVAE and dVAE respectively.

**Figure 4:**
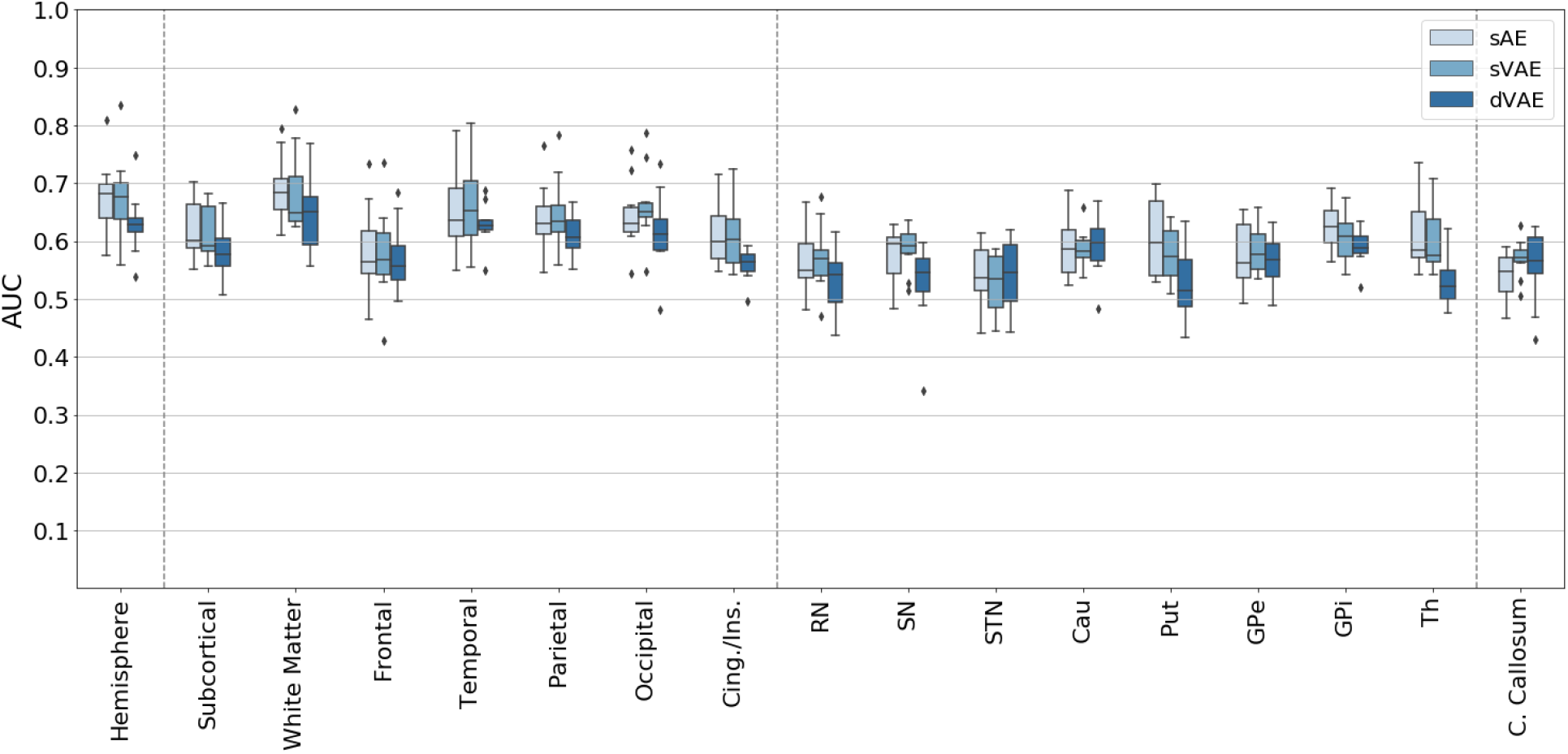
Obtained ROC-AUC scores for the whole brain and several ROIs using the sAE, sVAE and dVAE models. RN: Red Nucleus; SN: Substancia Negra; STN: Sub-thamic Nucleus, Cau: Caudate nucleus; GPe: Globus Pallidus external; GPi: Globus Pallidus internal; Th: Thalamus.

As explained in Section 4.2, each ROC curve provides a specific pathological threshold from which the g-mean score can be computed to assess how this ROI threshold differentiates controls from patients. For each considered ROI, the average over the 10 sub-populations (Table 1) of the ROC-AUC scores, of the retained pathological thresholds (p.t.) and of their associated g-mean scores provided by sAE are presented in Table 2. For a given ROI, the higher the p.t. the higher the percentage of abnormal voxels needs to be considered as pathological. A high pathological threshold indicates then that for HC in the test set, the auto-encoder has detected a relatively large number of abnormal voxels above the 2% expected errors. This means that HC in the test set exhibit more variability than the one captured by the network only built from HC in the learning set. For example, we expect that any hemisphere reconstructed with sAE with more than 2.18% of abnormal voxels, most likely belongs to a PD patient.

**Table 2:** AE reconstruction performance scores, expressed in percentages and averaged over the 10 sub-populations, for different ROIs (first column): ROC-AUC scores (2nd column), pathological thresholds retained from the ROC curves (3rd column) and associated g-mean scores (4th column).

| Structure | ROC-AUC | P. threshold | G-mean |
| --- | --- | --- | --- |
| Hemisphere | 67.6 | 2.18 | 64.6 |
| Subcortical | 62.0 | 1.02 | 61.2 |
| White Matter | 69.0 | 4.24 | 67.4 |
| Frontal | 58.4 | 1.02 | 60.4 |
| Temporal | 65.1 | 1.70 | 64.4 |
| Parietal | 64.7 | 0.80 | 61.1 |
| Occipital | 64.4 | 1.98 | 61.7 |
| Cing./Ins. | 61.0 | 1.66 | 58.5 |
| RN | 56.7 | 9.21 | 57.5 |
| SN | 57.6 | 20.96 | 57.7 |
| STN | 54.1 | 19.35 | 53.6 |
| Cau | 59.1 | 1.03 | 58.3 |
| Put | 60.5 | 1.08 | 60.4 |
| GPe | 57.6 | 2.91 | 57.6 |
| GPi | 62.5 | 5.11 | 62.2 |
| Th | 60.9 | 1.54 | 61.7 |

The g-mean classification scores for all models obtained for each ROI and each sub-population, are presented in Figure 5. We notice that the brain regions that obtained the highest scores for the sAE reconstructions are the White Matter and the Temporal lobe with a g-mean average of 67% (p.t. 4.24%) and 64% (p.t. 0.7%) respectively. For the subcortical structures, the internal part of the Globus Pallidus (GPi) and the Thalamus achieved the best performances in average with a g-mean score of 62% for both. However, the dispersion of the temporal lobe scores was quite important (standard deviation: 5.6%) compared to the other ROIs. When using the sVAE model, the g-mean score variation associated to the Temporal lobe was also the highest one out of all ROIs with a value of 65.1 ± 6.6%. For comparison, the White Matter obtained a g-mean score (sVAE) of 66.2 ± 4.3%, the GPi, 60.1 ± 3.5% and the Thalamus, 60.5 ± 3.2%.

**Figure 5:**
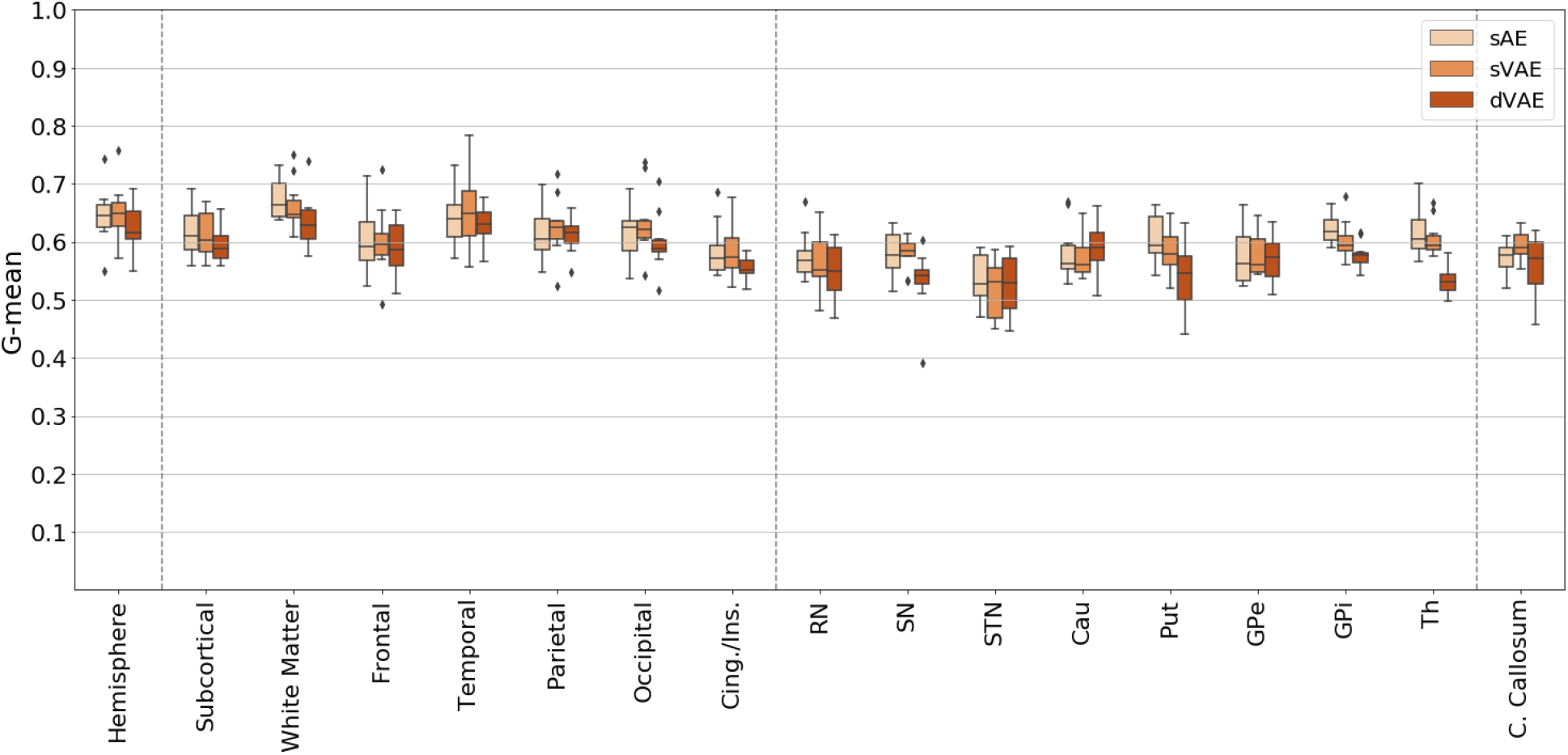
Obtained g-mean scores for the whole brain and several ROIs using the sAE, sVAE and dVAE models. For abbreviations see Fig. 4.

### 5.2. Sub-types of patients

According to the sub-typing criteria mentioned in Section 4.2, 34 patients were categorized as MCI, i.e. 68 hemispheres in our classification task. 66 patients had at least one hemisphere with a contralateral UPDRS III motor score above ten points. 59 had a dominant hemisphere and seven had bilateral motor disturbances. The classification procedure was repeated for these three subsets of patients compared to the ten previously defined control sub-populations using only sAE reconstructions. The gmean score results are displayed on Figure 6.

**Figure 6:**
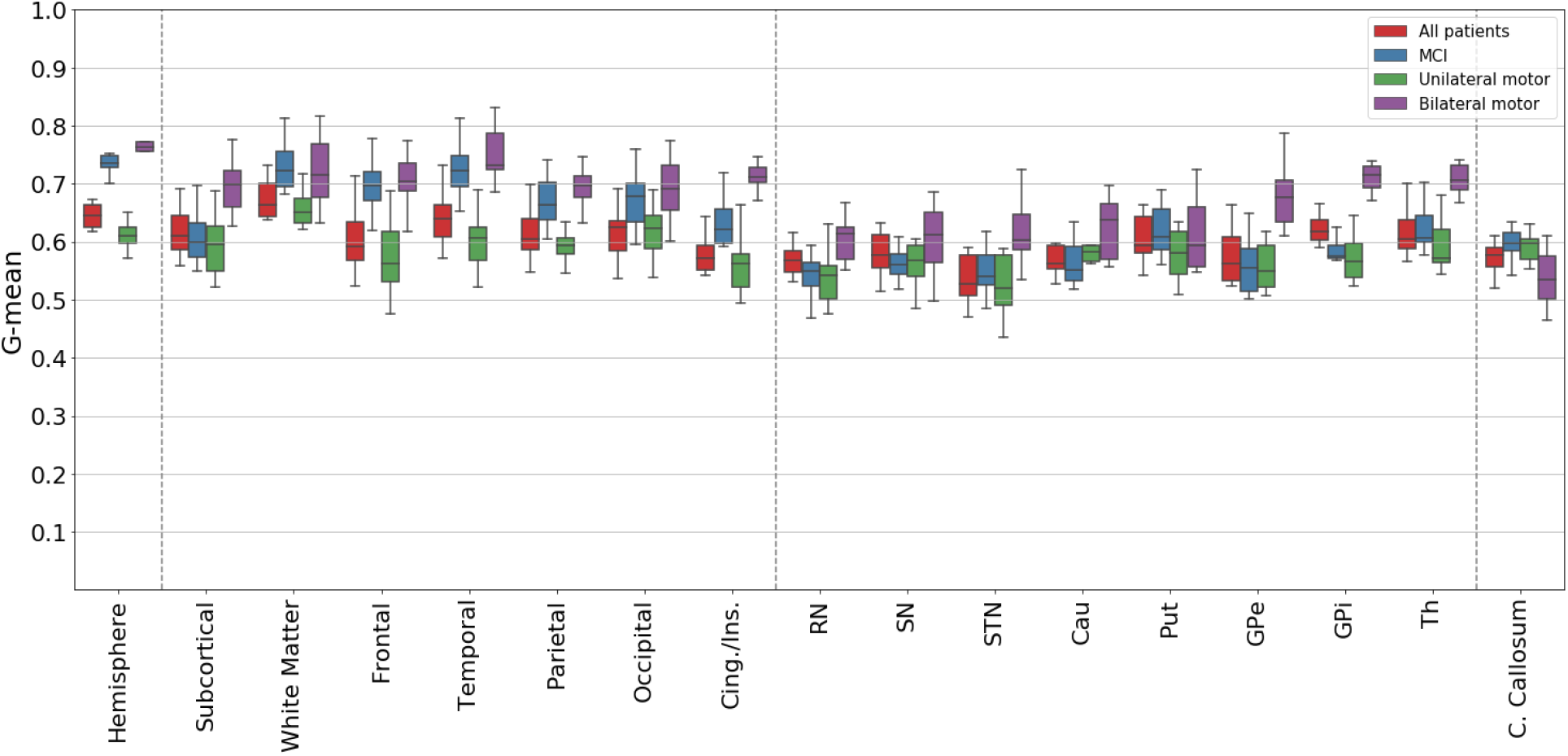
sAE reconstructions g-mean distributions for four classification tasks. Control hemispheres are compared to hemispheres respectively from: (red) the full patient population, (blue) the MCI subgroup (patients with MoCA score < 26), (green) patients with unilateral motor issues (UPDRS III score > 10 for only one hemisphere), and (purple) patients with bilateral motor issues (UPDRS III scores > 10 for both hemispheres).

Regarding the whole hemisphere classification, the hemispheres belonging to patients with bilateral motor symptoms obtained the highest g-mean scores with an average of 76.3% followed by hemispheres of patients with MCI (73.5%). The mean g-mean score for all hemispheres was of 64.6% and 63.1% for all contralateral hemispheres of motor symptoms. Hemispheres of patients with MCI had the best g-mean scores in average in the white matter (73%), the putamen (61.8%) and the corpus callosum (59.6%). The hemispheres belonging to patients with bilateral motor disturbances had the best gmean scores for the frontal (71.3%), temporal (74.9%), parietal (69.2%), occipital (69%) lobes and insular cortex (71.6%). This is also true for the association of subcortical structures (69.4%), with average of 61.2% for the RN, 60.6% for the SN, 61.5% for the STN, 62.3% for the caudate nucleus, 67.7% and 71.2% for the external and internal segment of the globus pallidus and 70.6% for the thalamus.

## 6. Discussion

Auto-encoders, and in general unsupervised deep anomaly detection techniques, are cost effective techniques that do not require annotated data for training. They are very promising for medical applications where annotated training examples are difficult to obtain. In this domain, they can learn the inherent features of a set of data, i.e. representative of the normal population, from which a patient can then be detected as an outlier. In this paper, we proposed to analyze MR brain scans with a deep learning anomaly detection technique, using either an auto-encoder or a variational auto-encoder as the main building block. We demonstrated that, when trained in an unsupervised manner with a medium-size collection of healthy MR brain scans, such a technique allowed the detection of subtle abnormalities in *de novo* PD diffusion feature maps.

An original evaluation procedure was designed to compare the reconstruction error profiles of healthy individuals versus PD patients. As a result, thresholds of abnormality were established for the whole brain and also for specific subcortical structures preferentially impacted by the disease. With an unbalanced dataset (56 controls vs 110 PD patients) we took special care in employing convenient metrics such as the geometric mean between specificity and sensibility, but also in generalizing our results by performing a 10-fold cross-validation to train and test our models. This turned out to be a key part in the study. As a matter of fact, for no obvious reason the first sub-population achieved much better performances than the average of our ten populations, with an sAE g-mean score associated to the whole brain of 74.3% when the average was 64.6% (see Figure 5).

While our results did not strongly highlight a particular biomarker for early PD, they provided an indication on the presence of diffuse anomalies mainly located in the white matter, all the more so in patients presenting cognitive decline. This white matter good performance is not very surprising considering that DTI has been introduced to study white matter through water molecules diffusion along the myelin fibers. In contrast, we suspect that the large pathological thresholds (see Table 2) obtained for small subcortical structures, like the substantia nigra (20.96%) and the STN (19.35%), are indicative that the spatial auto-encoder was unable to produce an accurate reconstruction of these regions in the HC population. The reasons for this could be related to the size of these structures and various registration issues induced by partial volume effects resulting in a somewhat bias control model. In that case, we may expect that the enlargement of the training healthy population would moderate such effects and allow to reinforce abnormalities detection in these subcortical structures for PD patients.

In terms of comparison with the literature, our approach achieved similar performances than other diffusion based studies. Notably, the cross-validation procedure of Schuff et al. (2015) obtained a ROC AUC of 59% for the rostral segment of the SN which is comparable to our ROC AUC of 57.6% for the complete SN (see Figure 4). In contrast, our results fall below those achieved by Li et al. (2019) employing a stacked spatial auto-encoder using morphometric (tissue concentration) and diffusion (mean diffusivity) features for 116 ROIs as input. Indeed they got a ROC AUC of 86% but they did not indicate any crossvalidation or generalization procedure. Still, it is possible that the relationship between white matter quantity, gray matter quantity and mean diffusion brings out anomalies that are not significant when studying diffusion properties alone.

Considering sub-types of patients based on their clinical scores, we found that patients with MCI were easier to identify. The classification g-mean scores of these patients hemispheres were better than those of the complete pool of patients when considering the whole hemisphere, the white matter and the gray matter lobes. In contrast, the classification performances for MCI patients were worse when focusing on subcortical structures, with the exception of the STN, the putamen and the thalamus. This could indicate a different type of diffusion anomalies in MCI patients.

The sub-typing of patients based on motor symptoms revealed that the hemispheres belonging to patients with bilateral motor symptoms were easier to differentiate from healthy hemispheres than hemispheres where motor symptoms were lateralized (see Figure 6). We note that this was the only classification task performed with more healthy samples (30) than patient samples (14). Also the average age of these patients (66.7 ± 9.2) was higher than those of controls (see Table 1). This could be at the root of the superior variance displayed in the results of ROIs like white matter and SN.

For the patients exhibiting lateralized motor symptoms, the classification scores achieved by their contralateral hemispheres were only superior in average to the results of the complete pool of patients for the occipital lobe, the caudate nucleus and the corpus callosum (see Figure 6). Interestingly, this latter structure was the only ROI providing superior classification score for patients with lateralized motor symptoms compared to bilateral motor symptoms. This is all in accordance with the literature. Indeed, while all the patients in the study are considered ‘de novo’, it is of common understanding that in the first stage of PD, motor symptoms are unilateral and only develop to bilateral disturbances over time (Hoehn and Yahr (1967)). What is more, the corpus callosum is constituted by white matter tracts that enable the communication between the left and the right hemispheres. Unilateral motor symptoms could then result from diffusion abnormalities in this structure.

In addition to the difficulty to interpret and relate our findings to the physio-pathology of the disease, the reliability of conclusions that can be drawn is subject to one of the major limitations of deep learning approaches. Indeed, the main challenge of applying deep learning to medical image analysis is data scarcity and this study is no exception. In face of the relative lack of available data for learning, the models were trained with 2D slices which multiplied by 40 the number of samples available for learning. The control group available for our experiments contained only 56 MRI volumes, with gender and age imbalance and thus not faithfully representing the variability of healthy brains in the population. Although we were able to discriminate between healthy controls and individuals affected by PD with good performances, we cannot rule out that other possible causes of variability in brain properties, such as age and gender, and other hidden parameters, might have influenced our classification results. Another inherent difficulty in neural networks implementation is the numerous tuning choices that can be made. While architecture-wise, reasonable settings seem to have emerged, it would worth investigating more sophisticated loss functions, e.g. with added adversarial components. More specific to anomaly detection, different anomaly scores could also be tested. At last, we cannot exclude limitations coming from imaging issues, such as the poor resolution of diffusion images. The atlas-based segmentation of anatomical regions may have consequently suffered from partial volume effect. This was probably negligible for large structures but might have had a significant impact on the results in small structures such as the SN, STN and RN.

Overall, more experiments, including a larger set of patients and controls and additional MR modalities, are required to confirm these preliminary and very encouraging results.

## 7. Conclusion

We developed an auto-encoder-based technique to detect subtle abnormalities in MRI brain scans of *de novo* and untreated patients with Parkinson’s disease. Based on FA and MD measures, we found the presence of diffuse structural anomalies, mainly located in white matter, especially in patients with cognitive disturbances. We may expect that the integration of additional quantitative MRI measures, such as perfusion, iron content and tissue relaxation times, would improve abnormalities detection in the brain of PD patients. Moreover, the approach could also be of interest for studying other neurological disorders when diffuse lesions are suspected (such as mild traumatic brain injury) and difficult to localize for a human observer. Moreover, the spatial localization of subtle alterations in MR imaging modalities, sensitive to different physiological parameters, could bring new knowledge about the physio-pathology of the underlying disease. Because they are sensitive to noise, corruption and overfitting, the main difficulty for disseminating deep learning methods in the medical domain is the requirement of a large amount of data. Our results offer compelling evidence that the deep anomaly detection technique employing auto-encoders could be used as a blueprint to detect subtle anomalies for medical purposes, even when trained with a moderate number of images.

## Data Availability

Data used in the preparation of this article is available at the Parkinsons Progression Markers Initiative (PPMI) database (www.ppmi-info.org/data).

## Acknowledgments

VMR is supported by a grant from NeuroCoG IDEX UGA in the framework of the “Investissements d’avenir” program (ANR-15-IDEX-02).

Data used in the preparation of this article were obtained from the Parkinson’s Progression Markers Initiative (PPMI) database (www.ppmi-info.org/data). For up-to-date information on the study, visit www.ppmi-info.org.

PPMI a public-private partnership is funded by the Michael J. Fox Foundation for Parkinson’s Research and funding partners, including Abbvie, Allergan, Avid Radiopharmaceuticals, Biogen, BioLegend, Bristol-Myers Squibb, Celgene, Denali, GE Healthcare, Genentech, GlaxoSmithKline, Lilly, Lundbeck, Merck, Meso Scale Discovery, Pfizer, Piramal, Prevail Therapeutics, Roche, Sanofi Genzyme, Servier, Takeda, Teva, UCB, Verily, Voyager Therapeutics and Golub Capital.

